# Evaluating a Motor Progression Connectivity Model Across Parkinson’s Disease Stages

**DOI:** 10.1101/2024.02.05.24302023

**Authors:** Mallory L. Hacker, David A. Isaacs, Nanditha Rajamani, Kian Pazira, Eli Abdou, Sheffield Sharp, Thomas L. Davis, Peter Hedera, Fenna T. Phibbs, David Charles, Andreas Horn

## Abstract

**Background:** Stimulation of a specific site in the dorsolateral subthalamic nucleus (STN) was recently associated with slower motor progression in Parkinson’s Disease (PD), based on the deep brain stimulation (DBS) in early-stage PD pilot trial.

**Objective:** To test whether stimulation of this site is associated with improvements of long-term motor outcomes in advanced-stage PD.

**Methods:** Active contacts of the early DBS cohort (N=14) were analyzed. Sweet spot and connectivity models derived from this cohort were then used to estimate long-term motor outcomes in an independent DBS cohort of advanced-stage PD patients (N=29).

**Results:** In early-stage PD, proximity of stimulation to the dorsolateral STN associated with slower motor progression. In advanced-stage PD, stimulation proximity to the same site associated with better long-term motor outcomes (R=0.60, P<0.001).

**Conclusions:** Results suggest stimulation of a specific site in the dorsolateral STN associates with both slower motor progression and long-term motor improvements in PD.

## INTRODUCTION

Recently, we demonstrated that deep brain stimulation (DBS) of a specific site within the dorsolateral region of the subthalamic nucleus (STN) is associated with slower motor progression for early-stage Parkinson’s disease (PD) patients^1^. This site receives cortical (hyperdirect) input from the primary motor (M1) and supplementary motor area (SMA) but not from the pre-SMA. These results built upon a post hoc analysis of the DBS for early-stage PD trial^2^. The specific site that we identified is similar to, yet slightly more ventral than, a previously reported metanalytic location associated with optimal symptomatic motor benefit in advanced-stage PD^3^. Moreover, a similar connectivity profile (i.e., stimulating M1/SMA) has also been associated with symptomatic motor improvement in advanced-stage PD^4–6^. Therefore, it could be that these sites, i.e., the one associated with slower motor progression and the one associated with optimal motor symptom improvements, are the same. However, such a relationship has not yet been empirically analyzed.

Our early-stage PD electrode localization study applied DBS sweet spot mapping^7^ and DBS fiber filtering^8^ to evaluate relationships between motor progression and stimulation location across the entire early DBS cohort^1^. This led to a specific site (sweet spot) and a set of connections associated with slower motor progression (connectivity model). Here, we analyze the relationship between active contacts and slower motor progression further, and we present subject-level visualizations of this early DBS dataset. We also evaluate whether the sweet spot and connectivity models constructed from the DBS in early-stage PD cohort could also explain significant amounts of variance in long-term motor improvements in an independent cohort of advanced-stage PD patients treated with DBS per standard of care.

## METHODS

### Cohorts

This study revisits data from Vanderbilt “DBS in early-stage PD” pilot clinical trial (NCT00282152; IDEG050016; IRB#040797), which was a prospective, randomized, single-blind clinical trial evaluating bilateral STN in patients with early-stage PD (aged 50-75 years; PD medication duration 1-4 years; no history or evidence of dyskinesia or motor fluctuations)^9^. All 14 subjects with complete data for the prior electrode localization study^1^ are included in this analysis, alongside clinical outcomes for subjects randomized to optimal drug therapy (ODT) in the pilot trial^9^. The present study also includes an independent cohort of 29 advanced-stage PD patients who received STN-DBS as standard care and participated in a long-term outcomes study at Vanderbilt University Medical Center (IRB#181198). The standard of care DBS subjects were recruited at least two years post-surgery. All subjects provided written informed consent for participation.

### Clinical Outcomes

In the early-stage PD cohort, motor progression was defined as the change in the Unified Parkinson’s Disease Rating Scale Part III score (UPDRS-III) from pre-operative baseline to 24 months measured after a seven-day washout (baseline: OFF Medications; 24 months: OFF Medications and OFF Stimulation), which was blindly rated from video-recorded motor examinations at the conclusion of the trial^9^. In the advanced-stage PD cohort, pre-operative (ON Medications) and longitudinal post-operative (ON Medications, ON Stimulation) UPDRS-III motor examinations were video-recorded and blindly rated. Long-term motor symptom changes were defined as the percent change from the pre-operative UPDRS-III to the longitudinal post-operative UPDRS-III. UPDRS-III scores for both cohorts do not include rigidity which cannot be evaluated via video. Levodopa equivalent daily doses (LEDD) were calculated for both cohorts as previously described^10^.

### Electrode Localizations

Electrodes for both cohorts were localized with Lead-DBS^11,12^ using pre-operative T1-weighted and T2-weighted magnetic resonance (MRI) scans and post-operative computed tomography (CT) scans. The same processing pipeline for localizations previously reported for the early DBS cohort^1^ was applied to the standard of care DBS cohort. Group visualizations were performed using Lead-Group^13^.

### Estimation of Stimulation Volumes and Validation of the Sweet Spot and Motor Progression Models

The motor progression sweet spot and connectivity models were defined using the early-stage PD cohort as previously published^1^. Using the early-stage PD sweet spot (defined by voxels covering by at least 3 Electric fields (i.e., E-fields) with a vector magnitude >0.2V/mm^1^), E-field magnitudes for the standard of care, advanced-stage PD cohort were Spearman rank-correlated with their long-term motor improvement. For the connectivity model, for each subject’s E-field, field magnitude values along each streamline that was defined within a normative structural connectome were denoted. These values were then Spearman rank correlated with motor progression scores to assign each streamline with a positive or negative correlation value (‘Fiber R-scores’). Together, streamlines defined the connectivity model. Critically, the model was used exactly as priorly published with all model parameters unchanged^1^. This model was used to estimate long-term motor improvement of subjects in the advanced-stage PD cohort. For each E-field in this independent cohort, overlaps with the connectivity model were calculated and average weighted by the E-field magnitudes (‘Weighted Means of Fiber R-scores’).

### Data Availability

The de-identified data from the standard of care DBS cohort will be made available upon reasonable request. The de-identified data and related study documents from the ‘DBS in early-stage PD’ trial are not being publicly shared at this time as they are currently being used for the development of a proprietary, multicenter, phase III, pivotal clinical trial (IDE G050016).

## RESULTS

### Early DBS Active Contacts Grouped by Motor Progression Thresholds

The early-stage PD cohort (13/14 male; mean disease duration 2.6 ± 1.9 years; 60.9 ± 6.9 years old) was randomized to surgery as part of the ‘DBS in early-stage PD’ pilot clinical trial^9^. Based on two-year motor progression scores, active contacts for the 14 early DBS patients were divided into two groups (top responding and remaining patients) and visualized. Since no objective rationale could define the threshold used for grouping, this was repeated at various thresholds. The active contact for the top responding subject localized to the dorsolateral STN (Fig. 1A). This patient’s UPDRS-III score in the DBS OFF and medication OFF state did not deteriorate two years after surgery. In fact, it became 2 points better. When lowering the threshold to include all subjects with improved two-year UPDRS-III OFF scores (n=4) into the group of ‘top responders’, their stimulation sites showed proximity around the same coordinate within the dorsolateral STN (Fig. 1B). Lowering the threshold further included subjects with increasingly more motor progression and showed increasing distance of active contacts to this stimulation site (Fig. 1C-1D). Fig. 1E-G shows motor progression, change in LEDD, and stimulation amplitude for the four top responding early DBS subjects featured in Fig. 1B compared to the remaining early DBS subjects and the subjects randomized to ODT. Early DBS subjects with active contacts outside of the dorsolateral STN (i.e., ‘typical responders’) had motor progression similar to the ODT control group (Fig. 1E). Early DBS subjects with active contacts close to the dorsolateral STN site required less medication on average at each follow-up visit as compared to baseline (Fig. 1F) and their mean stimulation voltages were lower than the typical DBS responders (Fig. 1G).

**Figure 1:**
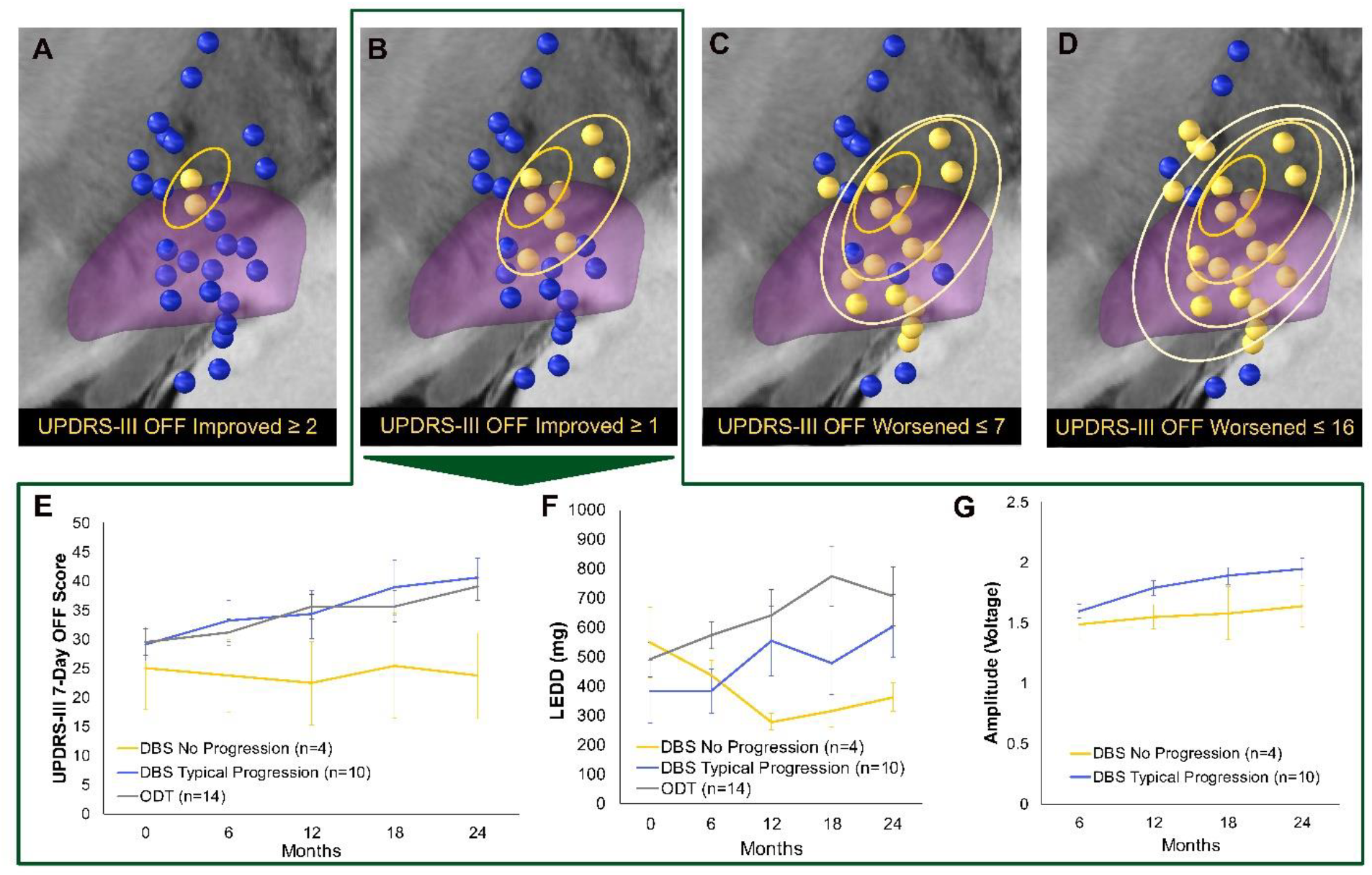
Active Contacts vs Motor Progression in the DBS in Early-Stage PD Pilot Clinical Trial Cohort. (A-D) Mirrored active contacts for the early DBS cohort are shown when dividing the cohort into two groups (yellow spheres = top responders; blue spheres = remaining subjects) at several motor progression thresholds (reported below each respective panel). (E-G) Clinical features of DBS subjects without motor progression whose active contacts are shown in panel B (yellow lines) vs the other DBS subjects (blue lines) and subjects randomized to the optimal drug therapy (ODT) control group (gray lines). Early DBS subjects without motor progression (A, yellow lines) required fewer PD medications (B) and lower stimulation amplitudes (C) Mean ± SEM.

### External Validation of the Motor Progression Sweet Spot and Connectivity Model

An independent cohort of 29 advanced-stage PD patients who received DBS as standard care (25/29 male; aged 60.1 ± 9.1 years at the time of surgery) was used to test the ability of the early-stage PD motor progression sweet spot and connectivity models^1^ to predict long-term motor improvements. Mean disease duration at the time of DBS surgery for this cohort was 9.9 ± 5.0 years, and mean time from surgery to post-operative follow-up was 5.4 ± 2.0 years. Long-term motor improvement was associated with optimal stimulation of the motor progression sweet spot in this independent cohort of advanced-stage PD patients (R=0.37, P=0.046; Fig. 2A-B). Optimal stimulation of the fiber tracts identified in the motor progression connectivity model also significantly associated with long-term motor improvement (P=0.60, P<0.001; Fig. 2C-D).

**Figure 2:**
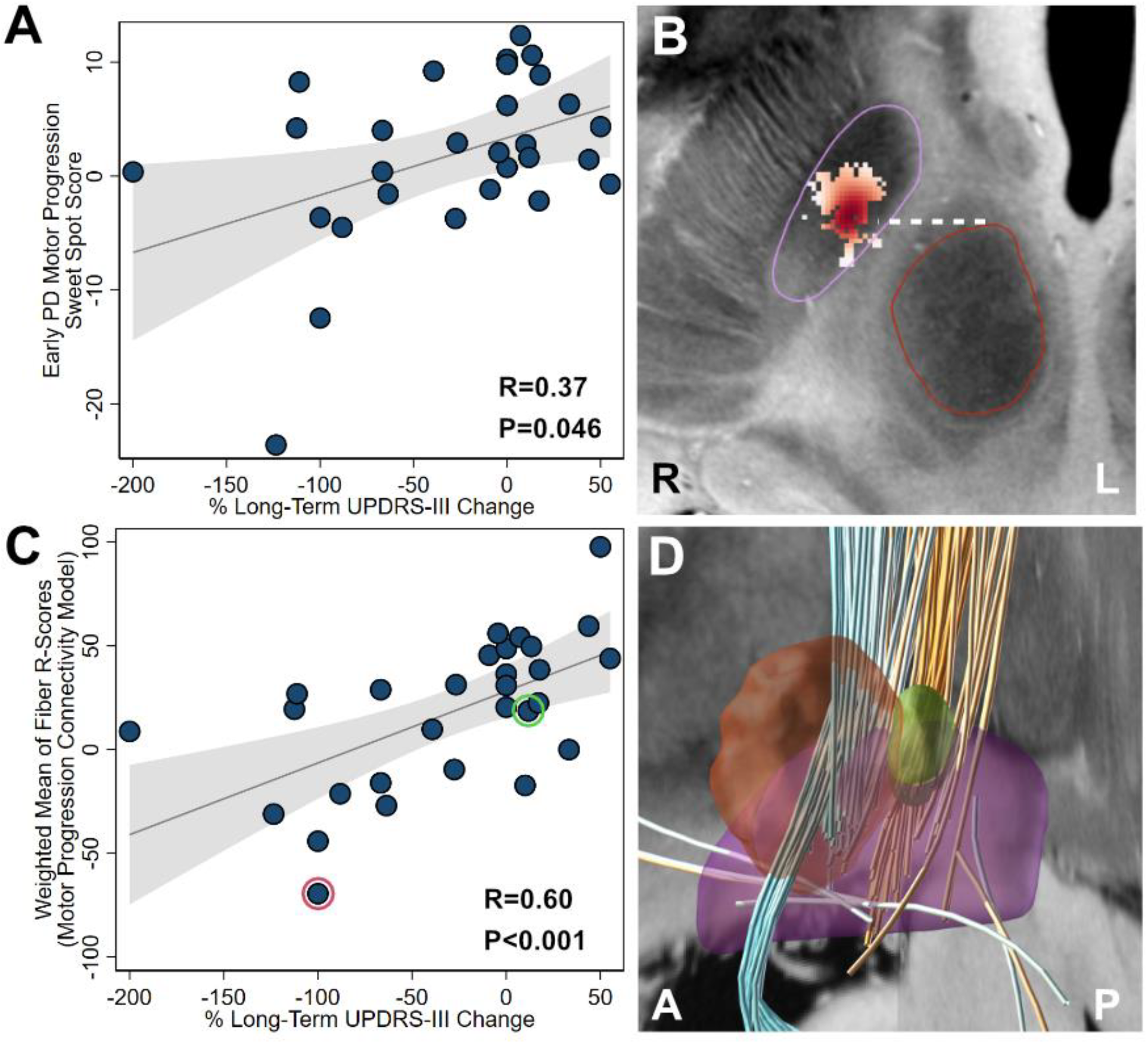
Validation of the Early PD Motor Progression Sweet Spot and Connectivity Model. The motor progression sweet spot (A-B) and connectivity model (C-D) identified in 14 early-stage PD patients^1^ were each used to estimate long-term motor outcomes in an independent cohort of 29 standard of care PD patients. (A) The degree of overlap with the motor progression sweet spot identified from the early DBS cohort (i.e., “Sweetspot Score”) significantly correlated with long-term UPDRS-III improvements (DBS duration at follow-up = 5.4 ± 2.0 years; Spearman’s R=0.37, P=0.046). (B) The motor progression sweet spot from^1^. STN is outlined in purple. Red nucleus is outlined in red. Bejanni line = white dashed line^15^. (C) The degree of stimulating positive tracts and not stimulating negative tracts identified from the early DBS cohort (i.e., the “Weighted Mean of Fiber R-Scores” of the Motor Progression Connectivity Model) significantly correlated with long-term UPDRS-III improvements (Spearman’s R=0.60, P<0.001). Two illustrative example patients are marked with colored circles in panel C and their stimulation volumes are shown in panel D (top responder example = green; poor responder example = red). (D) The motor progression connectivity model^1^ consists of positive (orange) tracts originating from M1 and SMA and negative (cyan) tracts originating from pre-SMA and cerebellum.

## DISCUSSION

There are three main findings in this study. First, we revisited the DBS in early-stage PD pilot clinical trial dataset to evaluate active contact locations at various thresholds to associate optimal stimulation location with the degree of motor progression on a single subject level. This confirmed that proximity to a specific site within the dorsolateral STN is associated with slower motor progression^1^. Second, motor progression in the patients from the early DBS trial that were stimulated outside of the dorsolateral STN was similar to motor progression in control subjects that received standard medical therapy in the trial. Finally, the motor progression sweet spot and connectivity models explained significant amounts of variance in long-term changes in motor symptoms in an independent group of advanced-stage PD patients.

Numerous studies have shown that stimulating a specific site within the dorsolateral STN associates with optimal motor improvement in advanced-stage PD^3,4,11^. The sites identified in these studies are remarkably similar to one another^14^ and seem to correspond to the same site associated with slower motor progression in our prior work^1^. Regarding brain connectivity, our previous study suggests that optimal site receives cortical input from M1 and SMA^1^. Critically, input from pre-SMA was instead negatively associated with slower motor progression. Here, we demonstrated that the same site – as identified based on slower motor progression – also accounted for long-term motor improvements in an independent typical advanced-stage (standard of care) PD cohort. This suggests that the site associated with slower motor progression is equally suited to maximize long-term motor improvements following STN-DBS for PD.

The identified optimal stimulation site is based on a low sample size, which is a key limitation. Unfortunately, additional cohorts that measure motor progression following DBS in humans do not yet exist, so the site may not be readily validated using additional data. However, very similar optimal stimulation sites have been described by others. Confirming utility of the stimulation site to estimate long-term motor improvements in an independent larger cohort may add further credibility.

## Conclusion

While the association of this dorsolateral STN stimulation site with slower motor progression needs to be further validated by additional prospective studies, our results explore the current data at hand and conclude that the same site is associated with both slower motor progression and long-term motor benefit with STN-DBS in Parkinson’s disease.

## Data Availability

The de-identified data from the standard of care DBS cohort will be made available upon reasonable request. The de-identified data and related study documents from the DBS in early-stage PD trial are not being publicly shared at this time as they are currently being used for the development of a proprietary, multicenter, phase III, pivotal clinical trial (IDE G050016).

## Notes

### Competing Interest Statement

Funding Sources and Conflict of Interest
The DBS in early-stage PD pilot trial was supported by Vanderbilt CTSA grants UL1TR000445 and UL1 TR002243 from the National Center for Advancing Translational Sciences (NCATS), NCATS/NIH award UL1TR000011, NIH R01EB006136, and Medtronic, Inc. The authors were free to independently design and conduct the study. Medtronic representatives did not take part in the collection, management, analysis, or interpretation of the data or in preparation, review, or approval of the manuscript. This work was also supported by a generous gift from Phyllis G. Heard, and her children, Elizabeth Heard, and Tony Heard.
MH receives funding from the National Institutes on Aging K01AG066971 and The Consolidated Anti-Aging Foundation. MH is a shareholder of Arena Therapeutics, a company focused on advancing research of DBS for the treatment of patients recently diagnosed with PD. DI receives funding from the National Institute of Neurological Disorders and Stroke (1K23NS131592) and from Teva Branded Pharmaceutical Products, R&D. DC is a shareholder of Arena Therapeutics, a company focused on advancing research of DBS for the treatment of patients recently diagnosed with PD. AH was supported by the German Research Foundation (Deutsche Forschungsgemeinschaft, 424778381 TRR 295), Deutsches Zentrum fur Luft- und Raumfahrt (DynaSti grant within the EU Joint Programme Neurodegenerative Disease Research, JPND), the National Institutes of Health (R01 13478451, 1R01NS127892-01, 2R01 MH113929 & UM1NS132358) as well as the New Venture Fund (FFOR Seed Grant). AH reports lecture fees for Boston Scientific and is a consultant for FxNeuromodulation and Abbott. There are no additional disclosures to report for NR, KP, EA, SS, FTP, PH, TLD.
Financial Disclosures for the previous 12 months:
MH receives funding from the National Institutes on Aging K01AG066971 and The Consolidated Anti-Aging Foundation. MH is a shareholder of Arena Therapeutics, a company focused on advancing research of DBS for the treatment of patients recently diagnosed with PD. DI receives funding from the National Institute of Neurological Disorders and Stroke (1K23NS131592) and from Teva Branded Pharmaceutical Products, R&D. DC is a shareholder of Arena Therapeutics, a company focused on advancing research of DBS for the treatment of patients recently diagnosed with PD. AH was supported by the German Research Foundation (Deutsche Forschungsgemeinschaft, 424778381 TRR 295), Deutsches Zentrum fur Luft- und Raumfahrt (DynaSti grant within the EU Joint Programme Neurodegenerative Disease Research, JPND), the National Institutes of Health (R01 13478451, 1R01NS127892-01, 2R01 MH113929 & UM1NS132358) as well as the New Venture Fund (FFOR Seed Grant). AH reports lecture fees for Boston Scientific and is a consultant for FxNeuromodulation and Abbott. There are no additional disclosures to report for NR, KP, EA, SS, FTP, PH, TLD.

### Funding Statement

The DBS in early-stage PD pilot trial was supported by Vanderbilt CTSA grants UL1TR000445 and UL1 TR002243 from the National Center for Advancing Translational Sciences (NCATS), NCATS/NIH award UL1TR000011, NIH R01EB006136, and Medtronic, Inc. The authors were free to independently design and conduct the study. Medtronic representatives did not take part in the collection, management, analysis, or interpretation of the data or in preparation, review, or approval of the manuscript. This work was also supported by a generous gift from Phyllis G. Heard, and her children, Elizabeth Heard, and Tony Heard.

### Author Declarations

The Human Protections Research Program of Vanderbilt University gave ethical approval for this work.

xI have followed all appropriate research reporting guidelines, such as any relevant EQUATOR Network research reporting checklist(s) and other pertinent material, if applicable.

## REFERENCES

1. Hacker, M. L., Rajamani, N., Neudorfer, C., Hollunder, B., Oxenford, S., Li, N., Sternberg, A. L., Davis, T. L., Konrad, P. E., Horn, A., & Charles, D. (2023). Connectivity Profile for Subthalamic Nucleus Deep Brain Stimulation in Early Stage Parkinson Disease. Annals of Neurology, 94(2), 271–284. 10.1002/ana.26674

2. Hacker, M. L., DeLong, M. R., Turchan, M., Heusinkveld, L. E., Ostrem, J. L., Molinari, A. L., Currie, A. D., Konrad, P. E., Davis, T. L., Phibbs, F. T., Hedera, P., Cannard, K. R., Drye, L. T., Sternberg, A. L., Shade, D. M., Tonascia, J., & Charles, D. (2018). Effects of deep brain stimulation on rest tremor progression in early stage Parkinson disease. Neurology, 91(5), e463–e471. 10.1212/WNL.0000000000005903

3. Caire, F., Ranoux, D., Guehl, D., Burbaud, P., & Cuny, E. (2013). A systematic review of studies on anatomical position of electrode contacts used for chronic subthalamic stimulation in Parkinson’s disease. Acta Neurochirurgica, 155(9), 1647–1654. 10.1007/s00701-013-1782-1

4. Akram, H., Georgiev, D., Mahlknecht, P., Hyam, J., Foltynie, T., Limousin, P., Jahanshahi, M., Hariz, M., Zrinzo, L., Ashburner, J., Behrens, T., Sotiropoulos, S. N., Jbabdi, S., & De Vita, E. (2017). Subthalamic deep brain stimulation sweet spots and hyperdirect cortical connectivity in Parkinson’s disease. NeuroImage, 158(January), 332–345. 10.1016/j.neuroimage.2017.07.012

5. Avecillas-Chasin, J. M., & Honey, C. R. (2020). Modulation of nigrofugal and pallidofugal pathways in deep brain stimulation for parkinson disease. Neurosurgery, 86(4), E387–E397. 10.1093/neuros/nyz544

6. Horn, A., Reich, M., Vorwerk, J., Li, N., Wenzel, G., Fang, Q., Schmitz-Hübsch, T., Nickl, R., Kupsch, A., Volkmann, J., Kühn, A. A., & Fox, M. D. (2017). Connectivity Predicts deep brain stimulation outcome in Parkinson disease. Annals of Neurology, 82(1), 67–78. 10.1002/ana.24974

7. Dembek, T. A., Baldermann, J. C., Petry-Schmelzer, J. N., Jergas, H., Treuer, H., Visser-Vandewalle, V., Dafsari, H. S., & Barbe, M. T. (2022). Sweetspot Mapping in Deep Brain Stimulation: Strengths and Limitations of Current Approaches. Neuromodulation, 25(6), 877–887. 10.1111/ner.13356

8. Baldermann, J. C., Melzer, C., Zapf, A., Kohl, S., Timmermann, L., Tittgemeyer, M., Huys, D., Visser-Vandewalle, V., Kühn, A. A., Horn, A., & Kuhn, J. (2019). Connectivity Profile Predictive of Effective Deep Brain Stimulation in Obsessive-Compulsive Disorder. Biological Psychiatry, 85(9), 735–743. 10.1016/j.biopsych.2018.12.019

9. Charles, D., Konrad, P. E., Neimat, J. S., Molinari, A. L., Tramontana, M. G., Finder, S. G., Gill, C. E., Bliton, M. J., Kao, C., Phibbs, F. T., Hedera, P., Salomon, R. M., Cannard, K. R., Wang, L., Song, Y., & Davis, T. L. (2014). Subthalamic nucleus deep brain stimulation in early stage Parkinson’s disease. Parkinsonism and Related Disorders, 20(7), 731–737. 10.1016/j.parkreldis.2014.03.019

10. Tomlinson, C. L., Stowe, R., Patel, S., Rick, C., Gray, R., & Clarke, C. E. (2010). Systematic review of levodopa does equivalency reporting in Parkinson’s disease. Movement Disorders, 25(15), 2649–2685. 10.1002/mds.23429

11. Horn, A., Li, N., Dembek, T. A., Kappel, A., Boulay, C., Ewert, S., Tietze, A., Husch, A., Perera, T., Neumann, W. J., Reisert, M., Si, H., Oostenveld, R., Rorden, C., Yeh, F. C., Fang, Q., Herrington, T. M., Vorwerk, J., & Kühn, A. A. (2019). Lead-DBS v2: Towards a comprehensive pipeline for deep brain stimulation imaging. NeuroImage, 184(August 2018), 293–316. 10.1016/j.neuroimage.2018.08.068

12. Neudorfer, C., Butenko, K., Oxenford, S., Rajamani, N., Achtzehn, J., Goede, L., Hollunder, B., Ríos, A. S., Hart, L., Tasserie, J., Fernando, K. B., Nguyen, T. A. K., Al-Fatly, B., Vissani, M., Fox, M., Richardson, R. M., van Rienen, U., Kühn, A. A., Husch, A. D., … Horn, A. (2023). Lead-DBS v3.0: Mapping deep brain stimulation effects to local anatomy and global networks. NeuroImage, 268(July 2022), 119862. 10.1016/j.neuroimage.2023.119862

13. Treu, S., Strange, B., Oxenford, S., Neumann, W. J., Kühn, A., Li, N., & Horn, A. (2020). Deep brain stimulation: Imaging on a group level. NeuroImage, 219(January), 117018. 10.1016/j.neuroimage.2020.117018

14. Horn, A. (2019). The impact of modern-day neuroimaging on the field of deep brain stimulation. Current Opinion in Neurology, 32(4), 511–520. 10.1097/WCO.0000000000000679

15. Bejjani, B. P., Dormont, D., Pidoux, B., Yelnik, J., Damier, P., Arnulf, I., Bonnet, A. M., Marsault, C., Agid, Y., Philippon, J., & Cornu, P. (2000). Bilateral subthalamic stimulation for Parkinson’s disease by using three-dimensional stereotactic magnetic resonance imaging and electrophysiological guidance. Journal of Neurosurgery, 92(4), 615–625. 10.3171/jns.2000.92.4.0615

